# Effectiveness of first line therapy for *Helicobacter pylori* infection in children and adolescents: a multicenter study in the United Arab Emirates

**DOI:** 10.1101/2025.01.16.25320652

**Authors:** Eman Al Atrash, Farah Al-Marzooq, Antoine AbdelMassih, Amina Bakro, Amer Azzaz

**Affiliations:** Division of Pediatric Gastroenterology, Department of General Pediatric, Sheikh Khalifa Medical City, Abu Dhabi, United Arab Emirates; Departments of Medical Microbiology and Immunology, College of Medicine and Health Sciences, United Arab Emirates University, Al Ain, United Arab Emirates; Pediatric Cardiology Division, Cardiac Sciences Department, Sheikh Khalifa Medical City, Abu Dhabi, United Arab Emirates; Division of Pediatric Nephrology, Department of General Pediatric, Sheikh Khalifa Medical City, Abu Dhabi, United Arab Emirates

## Abstract

Globally, gastric infection with *Helicobacter pylori* has a high prevalence. Bacterial eradication is crucial to prevent long-term complications such as gastric cancer. There is a standard recommended first-line therapy with a desirable eradication goal of 90%, but this is non-achievable in most trials in children. Thus, it is recommended to evaluate first-line therapy at national centres, especially in areas where antimicrobial susceptibility testing is limited. This study aimed to examine the effectiveness of first-line regimen in eradicating *H. pylori* infection in the pediatric population in the United Arab Emirates. This retrospective study was conducted in 2017-2023, involving patients aged 1-16 years. Patients diagnosed with and treated for *H. pylori* infection were included, specifically those who completed 2-week treatment with documented adequate compliance. Confirmation of *H. pylori* eradication was done 4 weeks after the completion of antimicrobial therapy. *H. pylori* was detected among 447 patients, while 413 patients underwent complete treatment. Only 345 patients (77%) were evaluated for confirmed eradication via stool *H. pylori* antigen test (44%) or urea breath test (39%) which were the most used diagnostic modalities. Most patients (285/345; 83%) were treated with Proton Pump Inhibitor/Amoxicillin/Clarithromycin (PAC). Eradication was successful in 226 patients (65%), mostly in patients treated with the PAC regimen (193/285; 68%), compared to 20-55% eradication rates for patients treated with other regimens including metronidazole. Antibiotic regimen was the only significant predictor of eradication success (p<0.001), with non-significant correlation with other factors as age and gender. The overall eradication rate for the first-line therapy for *H. pylori* falls below the recommended primary success rate for eradication (>90%). Factors influencing treatment success and adherence should be carefully considered to optimize the management of *H. pylori* infection in pediatric patients. Further prospective studies are warranted to validate these findings and explore additional strategies for improving treatment outcomes.

## Introduction

*Helicobacter pylori* is a Gram-negative, flagellated, helical bacterium with rising global prevalence [1]. It is estimated that approximately half of the world’s population have *H. pylori* infection or at least asymptomatic colonization in the stomach [2,3]. It has been designated a Class I carcinogen by the World Health Organization (WHO)[4]. Infection with *H. pylori* is a recognized risk factor for the emergence of gastric ulcers and gastric carcinoma. Almost 90% of all gastric malignancies are correlated to *H. pylori* infection. Socioeconomic status and hygiene levels affect the spread and prevalence of *H. pylori* infection; therefore, it is more common in developing than developed countries [5]. The prevalence of *H. pylori* infection varies widely with geographic locations, ranging between 85-95% in developing countries and 30-50% in developed countries [6,7]. Infection rate with *H. pylori* was found to be 20% among adolescents in the United States compared to >50% by 5 years of age and >90% in the adulthood in the developing world [5]. Among children, it is estimated at 32-36% worldwide [3]. Limited data is available from the United Arab Emirates (UAE), and the Middle East in general [8].

Prevalence in the UAE is thought to be much higher than suggested by the available recent data, underestimating local prevalence of *H. pylori* infection to a rate of 41% among the multinational general population and 55% among native Emiratis [8]. A few studies from the UAE in the past three decades were non-representative of the general population and restricted to high-risk groups [1,8,9]. Prevalence in UAE is expected to match that of similar nations of the Middle East and North Africa region where the average prevalence is assessed to be 72% [10]. A previous study from the UAE delineated a significant association between *H. pylori* infection, gender, age, ethnicity, profession, domestic overcrowding, source of drinking water, and gastrointestinal characteristics of participants [8].

*H. pylori* eradication is considered challenging. It can cure gastritis and can alter the progression to long-term complications. Guidelines on the management of *H. pylori* infection recommend eradication by administering two antimicrobials and a proton pump inhibitor (PPI), and/or a bismuth salt for 10-14 days [11]. Most used antibiotics are amoxicillin, clarithromycin and metronidazole. Other antibiotics used are levofloxacin, tetracycline and rifampicin [11]. It is recommended that eradication therapies must be tailored according to the local antimicrobial sensitivity patterns for the infecting *H. pylori* strains whenever possible [12]. The desirable goal of these therapies is at least 90% eradication rate [12].

Based on the limited data on the local *H. pylori* antimicrobial susceptibility patterns in the UAE and the lack of such service for routine clinical practice and according to the joint European Society for Pediatric Gastroenterology Hepatology and Nutrition (ESPGHAN) and North American Society for Pediatric Gastroenterology, Hepatology and Nutrition (NASPGHAN) guidelines for the management of *H. pylori* in children and adolescents which is updated in 2016 [12], it is proposed that national/regional effectiveness of first-line eradication regimens must be evaluated. There are no UAE studies evaluating the efficacy of *H. pylori* eradication regimens in children and adolescents. With the proposition that our patient population has a high treatment failure rate to first-line therapy, this study aims to analyse local treatment outcomes of *H. pylori* infection and to identify factors associated with such outcome in paediatric and adolescent population in the UAE.

## Materials and Methods

### Ethical Approval

The study received approval from the Institutional Review Board (IRB) of Sheikh Khalifa Medical City and Tawam Hospital (REC-22.05.2023 [RS-793]). Due to the retrospective nature of the study, patient consent for the review of their medical records was not required by the IRB. Patient data confidentiality was strictly maintained at every stage of the study, by deidentification of data and secured data sharing.

### Study Population

This is a retrospective multicentre observational study of pediatric patients aged 1-16 years old, that were looked after in Sheikh Khalifa Medical City, Abu Dhabi and Tawam Hospital, Al-Ain between January 1, 2017 to December 31, 2023, with proven diagnosis of *H. pylori* infection based on International Classification Diseases ICD Ninth and Tenth Revisions.Authors doesn’t have access to information that could identify individual participants during or after data collection.

Inclusion criteria were patients with documented *H. pylori* infection undergoing treatment for 14 days followed by assessment of *H. pylori* eradication confirmation 6-8 weeks after completion of therapy. In addition, patients needed to be off antibiotics and proton pump inhibitos for at least 4 and 2 weeks respectively when the *H. pylori* eradication confirmation test was performed. All patients whether *H. pylori* infection naïve or with history of *H. pylori* infection were included.

*H. pylori* infected patients with no records of eradication therapy or received treatment less than 2 weeks only and those who did not undergo eradication confirmation testing under standard conditions after completion of eradication therapy were excluded from *H. pylori* eradication assessment analysis. Adequate compliance was defined as the completion of the total 2 weeks of treatment documented in clinical record.

### Data collection and diagnostic tests used in the study

The data on the patients’ demographic characteristics, indications for *H. pylori* testing, diagnostic modalities performed, type and doses of antibiotic regimen used, and eradication test results were collected from each patients’ electronic medical records.

Diagnosis of *H. pylori* infection among patients is based on the results of one or more of the following tests; upper GI endoscopy with either biopsy showing *H. pylori* on histopathological examination or Campylobacter-like organism (CLO) test or both. Other investigative modalities including *H. pylori* stool antigen test (SAT) by enzyme immunoassay technique and ^13^C urea breath test (^13^C-UBT) were taken into account in assessment of *H. pylori* infection eradication [13,14].

### Statistical Analysis

Data were analyzed using Medcalc statistical software (Medcalc ltd 2023-Trial version) and Excel (Microsoft ltd). Numerical data were expressed in terms of mean±SD, while categorical data were expressed as percentages. The study subjects were then divided into two groups according to eradication success, numerical data were compared across groups using T-test while categorical data were compared using Chi-square test and Fisher test, when proportion in one group is zero. A multivariate regression was conducted to determine the best predictor of eradication, and a ROC analysis was implemented to determine the sensitivity of the latter to predict eradication. A *P* value <0.05 was considered statistically significant.

## Results

The sampling strategy and study population overview are illustrated in **Fig 1**.

**Fig 1.**
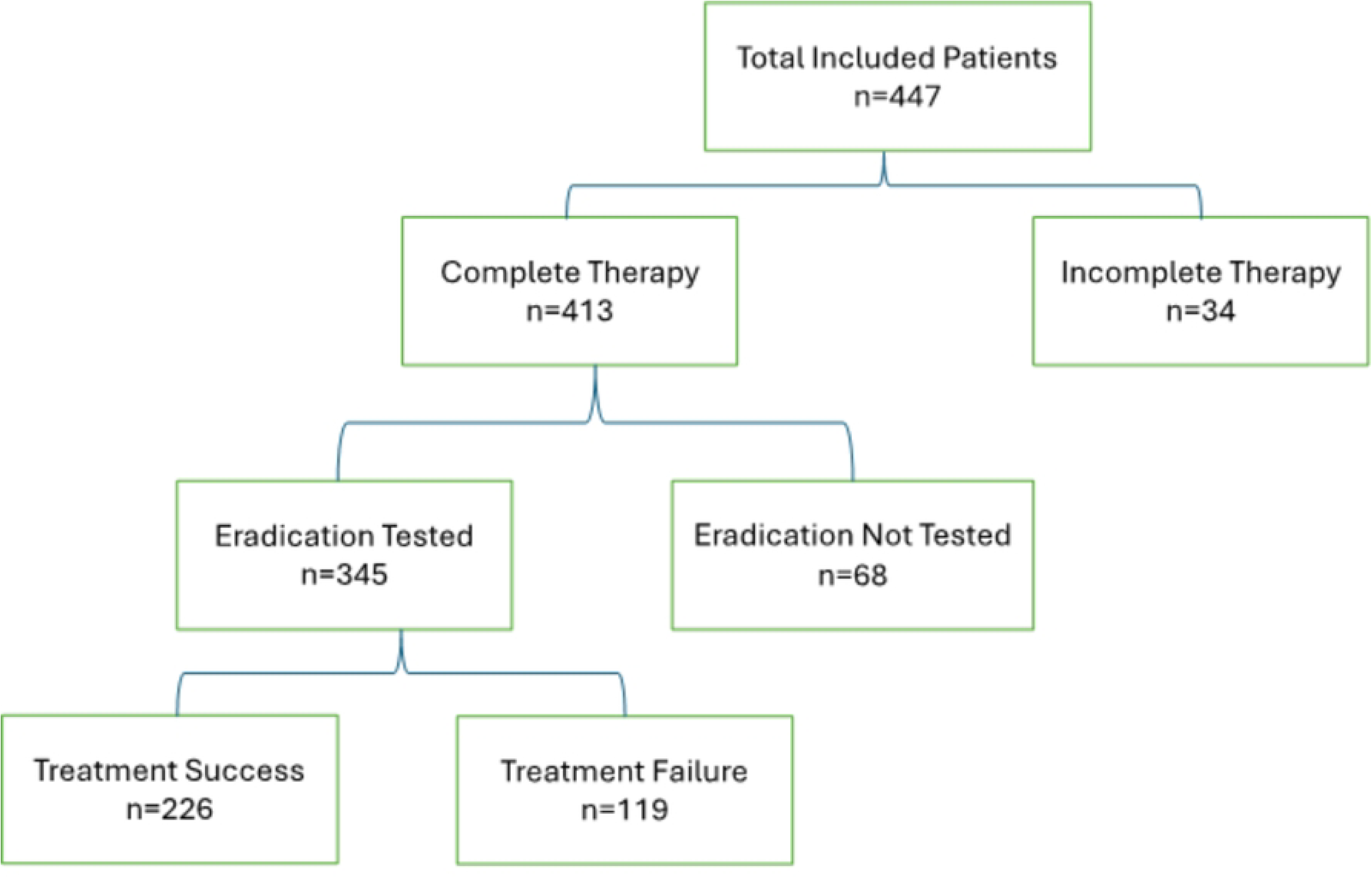
Overview of the study population, and steps for selecting patients for inclusion in the study.

A total of 447 patients infected with *H. pylori* were identified. Out of these, 413 patients underwent complete treatment for *H. pylori* infection. Treatment outcomes were analysed in 345 patients (77.0%), who had confirmed eradication via stool *H. pylori* antigen, EGD with biopsy, or urea breath test.

Stool *H. pylori* (196/447; 44%) and urea breath test (176/447; 39%) were the predominant diagnostic modalities. The mean age of the study population was 10 years (SD=3.54, range: 1-16 years old), with females accounting 58% of the total population. The demographic and clinical characteristics of the study population are summarized in **Table 1**.

**Table 1.** Demographic and clinical characteristics of the study population.

| Variables | N (%) |
| --- | --- |
| **Gender** |  |
| Male | 185 (41.4) |
| Female | 262 (58.6) |
| **Age groups (years)** |  |
| 1-5 | 63 (14.1) |
| 6-11 | 200 (44.7) |
| 12-16 | 184 (41.2) |
| **Reason for testing for *H. pylori*** |  |
| Abdominal pain | 379 (85) |
| Incidental finding | 18 (4) |
| Weight loss | 13 (3) |
| Contact with Infected Person | 9 (2) |
| Vomiting | 9 (2) |
| Dysphagia | 5 (1) |
| Diarrhea | 6 (1) |
| Heartburn | 4 (1) |
| Hematemesis | 2 (0.5) |
| Fecal donor assessment | 2 (0.5) |
| **Methods used to confirm eradication** |  |
| Fecal *H. pylori* antigen test | 183 (53) |
| Urea breath test | 147 (43) |
| Histology and CLO test | 15 (4) |
| **Utilized treatment regimens** |  |
| Proton Pump Inhibitor/Amoxicillin/Clarithromycin (PAC) | 285 (83) |
| Proton Pump Inhibitor/Amoxicillin/Metronidazole (PAM) | 36 (10) |
| Proton Pump Inhibitor/Clarithromycin/ Metronidazole | 10 (3) |
| Proton Pump Inhibitor/Amoxicillin/Clarithromycin/ Metronidazole (PACM) | 10 (3) |
| Bismuth-based regimen | 4 (1) |

Symptoms related to the gastrointestinal (GI) tract prompted *H. pylori* testing in 400/447 cases (89%), primarily driven by abdominal pain (84%), other symptoms include vomiting, hematemesis, heartburn, diarrhea, and dysphagia in total of 5% of the study population. One patient exhibited major upper gastrointestinal bleeding characterized by hematemesis (Table 1).

Additional reasons for screening were initiated due to incidental finding during elective endoscopy in 5% of the cases, contact with diagnosed individuals in 2%, and elective assessments for fecal donors in 1%. (**Table 1**).

Among 77 patients who underwent upper esophagogastroduodenoscopy (EGD), 45% exhibited normal EGD findings. Gastric mucosa erythema was detected in 14% of cases. Gastric nodularity without gastric erythema was observed in 24%, while 12% displayed both gastric erythema and nodularity, contrasting with the 5% who exhibited the erythematous gastric mucosa and nodularity alongside gastric ulcers. Most patients (80%) had normal macroscopic findings in duodenum, while 10% of the patients had duodenal ulcer. Only one patient had active bleeding during endoscopy.

Most patients (300/345; 90%) received antibiotic for ≥14 days and 85% of patient received at least 2 weeks of proton pump inhibitor. Esomeprazole was the most used PPI in 54% of the patients. Treatment outcomes were assessed using fecal *H. pylori* antigen test (183; 53%), urea breath test (147; 43%), or histology and CLO test (15; 4%).

The most utilized treatment regimen was Proton Pump Inhibitor/Amoxicillin/Clarithromycin (PAC) in 285/345 patients (83%), followed by Proton Pump Inhibitor/Amoxicillin/Metronidazole (PAM) in 36/345 patients (10%), Proton Pump Inhibitor/Amoxicillin/Clarithromycin/Metronidazole (PACM) and Proton Pump Inhibitor/Clarithromycin/Metronidazole were equally used in 10 patients for each accounting for 3% (10/345), while bismuth-based regimen was used in 1 % (4/345).

Overall eradication was successful in 226 patients (65%). Among those treated with the PAC regimen, eradication was achieved in 193 out of 285 patients (68%), compared to 20 out of 36 patients (55%) with PAM. The least successful eradication rate was with proton pump inhibitor/Clarithromycin/Metronidazole regimen at 20% (*p*<0.009) as shown in **Fig 2**.

**Fig 2.**
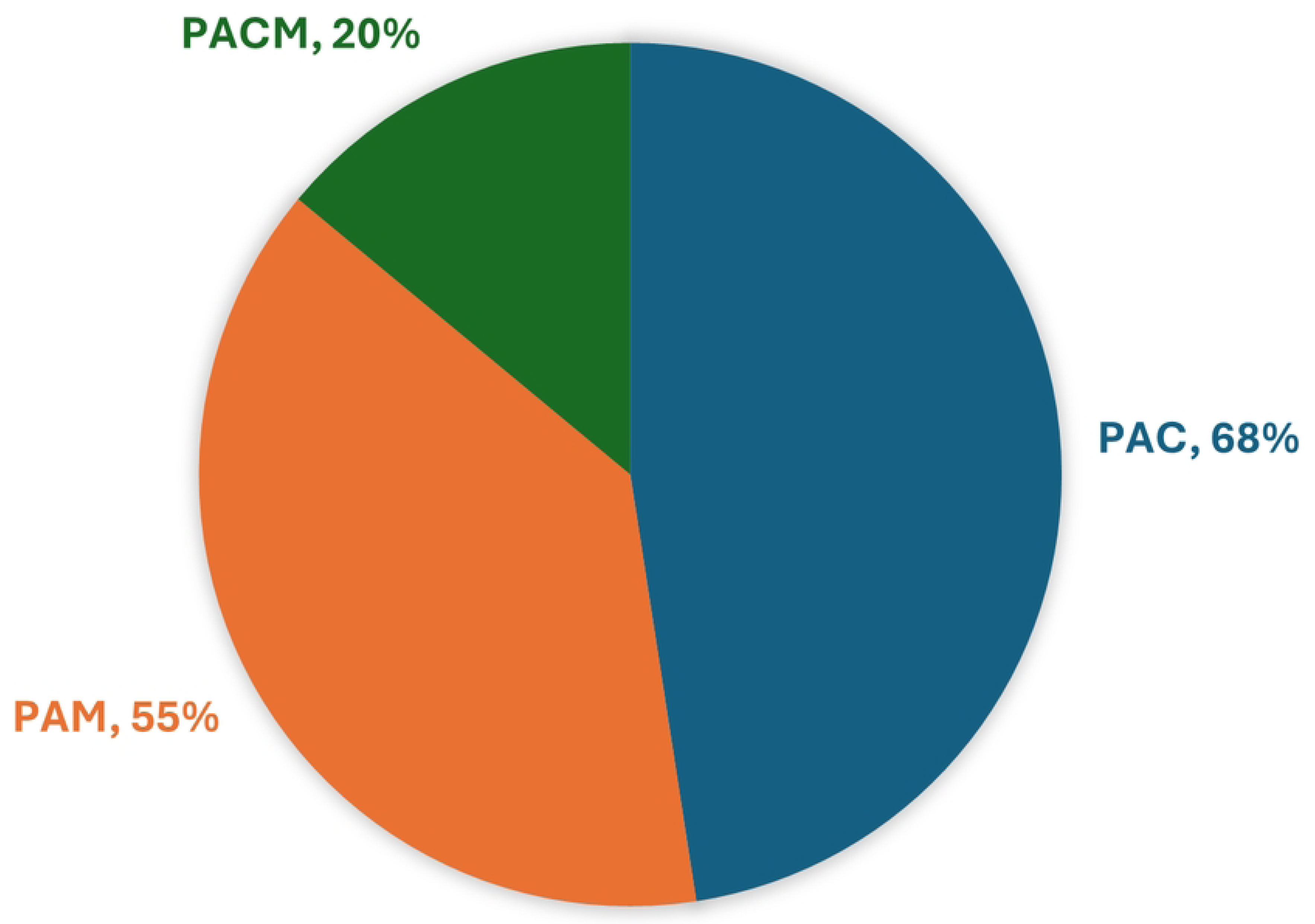
Eradication success rate for each antibiotic regimen used. PAC: Proton pump inhibitor, amoxicillin, and clarithromycin. PAM: Proton pump inhibitor, amoxicillin, and metronidazole. PAMC: Proton pump inhibitor, amoxicillin, metronidazole and clarithromycin.

Multivariate analysis showed that age (*p*=0.99), gender (p=0.098), type of proton pump inhibitor used (*p*=0.24), duration of proton pump inhibitor (*p*=0.97) and duration of antimicrobial treatment (*p*=0.06) were not associated with eradication success rate (**Table 2**). Antibiotic regimen is the only significant predictor of eradication success with *p*<0.001.

**Table 2.** Multivariate analysis for prediction of eradication success (* significant difference)

| Independent Variables | Coefficient | *P* value |
| --- | --- | --- |
| Age | -0.00008925 | 0.9919 |
| Gender | 0.09033 | 0.0984 |
| Antimicrobial regimen | -0.1976 | **<0.0001\*** |
| Antimicrobial duration | -0.1288 | 0.6317 |
| PPI type | 0.06286 | 0.2418 |
| Amoxicillin Standard Dose | 0.02784 | 0.6552 |
| Clarithromycin Standard Dose | -0.06903 | 0.3629 |

Among the 253 patients who received the standard amoxicillin dose recommended by the Joint ESPGHAN/NASPGHAN Guidelines for the Management of *Helicobacter pylori* in Children and Adolescents (Update 2016) as shown in Table 3 [12], only 171 patients had eradication success (*p*=0.5). High dose of amoxicillin in combination with clarithromycin and proton pump inhibitor is recommended if sensitivity was unknown, thus, it was used in 70 patients. Eradication success was achieved in 27% of these patients with no clinically significant difference compared to patient who received the same regimen with the standard amoxicillin dose (*p*=0.1).

**Table 3.** Standard dosing regimen recommended by the Joint ESPGHAN/NASPGHAN Guidelines for the Management of *Helicobacter pylori* in Children and Adolescents (Update 2016)

| Drug | Bodyweight Range<br>(kg) | Dose (mg) |  |
| --- | --- | --- | --- |
|  |  | Morning | Evening |
| Amoxicillin | 15-24 | 500 | 500 |
|  | 25-34 | 750 | 750 |
|  | >35 | 1000 | 1000 |
| Clarithromycin | 15-24 | 250 | 250 |
|  | 25-34 | 500 | 250 |
|  | >35 | 500 | 500 |

Additionally, 253 patients received the standard recommended clarithromycin dose. Eradication success was reported in 171/253 (68%) with no significant correlation with eradication success (*p*=0.44).

## Discussion

*H. pylori* infection is one of the most common illnesses worldwide, and one of the most widespread childhood infections [15]. Several studies emphasized the importance of timely eradication of *H. pylori* to prevent peptic ulcer and gastric cancer [16], however, there is a significant knowledge gap on the effectiveness of eradication therapy, especially in the pediatric population. This retrospective multicentre study describes the efficacy of first-line therapy for *H. pylori* infection among children and adolescents in the UAE. This study is the first of its kind in the UAE, for several reasons. Firstly, it is the only multicentre study, with large sample size. Secondly, the limited studies from the UAE focused mainly on adult patients, and no studies targeted the paediatric population. Lastly, previous studies focused on microbiological properties, or seroprevalence, without considering the clinical outcomes and response to therapy [8,1,17].

The findings of this study are alarming, as it shows early infection with *H. pylori* at a very young age (as low as 1-year old), in line with studies from other parts of the world, with reports for early detection at very young ages in counties like Libya, Bangladesh, India [5], China, Taiwan, Japan and South Africa [18]. This is a serious public health issue as *H. pylori* infection is associated with the development of a variety of gastrointestinal and extra-gastrointestinal disorders. Microscopic gastritis is a constant manifestation, usually linked with 1-10% risk of developing gastric or duodenal ulcers, 0.1-3% risk of developing gastric adenocarcinoma, and <0.01% of developing mucosa-associated lymphoid tissue (MALT) lymphoma [18,19]. Numerous studies have indicated that eliminating *H. pylori* infection reduces the likelihood of peptic ulcer recurrence and further diminishes the risk of developing gastric cancer [20]. In contrary, delaying the eradication of *H. pylori* has been associated with an increased risk of peptic ulcer recurrence and the onset of gastric malignancies [21,22]. Furthermore, early infections in childhood are postulated to induce a low-grade chronic inflammation with progression into pre-malignant changes and eventually gastric carcinoma. Besides, pathologic evidence suggests that early infections with *H. pylori* can cause rapidly destructive gastric mucosal lesions, which could conceivably damage the gastric acid barrier even during childhood [18]. Therefore, it is advisable to promptly and effectively treat and eliminate *H. pylori* infection to prevent the occurrence of peptic ulcers and associated complications. *H. pylori* is usually transmitted in childhood and persists for life if untreated [14].

The high prevalence of this bacteria in early childhood and adolescent as seen in this study, is very concerning and require special attention to screening for early diagnosis. The pathophysiology of *H. pylori* infection relies on complex interaction of bacterial virulence, host immune system and environmental factors, resulting in distinct disease phenotypes that determine possible progression to different GI pathologies [23]. This was also reflected in this study, as patients present with various complains, suggesting variations in pathogenesis. A previous study in the UAE showed high prevalence of bacterial virulence factors such as Cytotoxin-Associated Gene (Cag A). Notably, 76.11 % of Emirati adult participants with *H. pylori* infection were positive for CagA [17]. This underscores the significance of this infection and emergence of strains with potent pathogenic potential, that may lead to serious complications as seen in other studies [24].

The efficacy of the *H. pylori* eradication therapy has decreased dramatically in most of the regions monitored by the WHO because of the rapid increase in the prevalence antibiotic resistant strains [24,25]. With rare exceptions, clarithromycin-containing regimens are considered no longer suitable for unconditional empiric use because of inadequate eradication rates of <80% [26]. Despite the international consensus reports strongly recommending the selection of treatment based on local resistance patterns, *H. pylori* antibiotic susceptibility testing is rarely performed [27]. Among the 45 countries investigated in the most recent global study of the prevalence of antibiotic resistance in *H. pylori* in 2018, there were no informative data from the UAE [3]. However, considering 15% as the threshold for high resistant rate as per the most recent international guidelines, the neighbouring countries of the East Mediterranean Region ranked the highest in pooled prevalence of *H. pylori* resistance to clarithromycin, metronidazole and levofloxacin with rates up to 57%, 74%, and 46%, respectively [28]. Resistance to amoxicillin and tetracycline remained ≤10% in all WHO regions [28]. In 2009, a small sample of 26 patients from UAE with *H. pylori* gastritis were investigated using PCR molecular testing and reported resistance to clarithromycin and metronidazole in 9 (35%) and 3 (12%) of the strains, respectively [29]. Another study described 65.5% (36/55) resistance to clarithromycin by PCR gene mutation testing and sequencing for the 23S rRNA gene and an additional one of 19.2% (5/26) based on traditional culture and antimicrobial susceptibility [30].

Amoxicillin with clarithromycin and a proton pump inhibitor is one of the preferred regimens to eradicate *H. pylori* in both adults and children. Therapeutic trials in children with this regimen yield success rates between 58-92% [18]. This is in accordance with our study, as the most effective therapeutic regimen (effective in 68% of the cases) was the combination of proton pump inhibitor, amoxicillin, and clarithromycin (PAC). This regimen was the most used in the study population in 285 (83%) patients. Combination of proton pump inhibitor, amoxicillin, and metronidazole (PAM) was also highly effective in 55% of the cases, although it was used in less patients (36; 10%). Other therapeutic regimens were less used and were remarkably less effective as combination of proton pump inhibitor, amoxicillin, metronidazole and clarithromycin (PACM) which achieved 20% effectiveness, when used in 10 (3%) patients. Our results contradict with the findings from a randomized controlled trial in Japan, which found that metronidazole-based triple therapy (PAM) achieved significantly higher eradication rates (98.3-100%) compared to clarithromycin-based therapy (PAC) with eradication rates of 60.5-63.4% in adolescents, suggesting PAM as a more effective first-line treatment when antibiotic susceptibility tests are unavailable [31]. However, another randomized trial in Taiwan showed that esomeprazole and amoxicillin-containing high-dose dual therapy regimen achieves a high eradication rate as first-line anti-*H pylori* treatment [32].

The increasing challenge of antibiotic resistance in treating *H. pylori* underscores the need for updated guidelines by NASPGHAN and ESPGHAN [12]. These guidelines apply only to pediatric patients, defined as children and adolescents below 18 years of age. Recommendations for children and adolescents may differ from recent guidelines for adults because of a different risk-benefit ratio depending on age and the fact that some antibiotics are not released or licensed for the pediatric population. Given the insufficient pediatric data across various regions worldwide, these guidelines suggest assessing eradication rates in different geographical areas.

In our study population from 2017 to 2023, the antibiotic combination used most frequently in the study was amoxicillin and clarithromycin, followed by amoxicillin and metronidazole. These treatment regimens are not in line with the last NASPGHAN and ESPGHAN guidelines of 2016, which recommends against using clarithromycin-based regimen if sensitivity is unknown.

The consensus group recommended that invasive diagnostic testing for *H. pylori* be performed only when treatment will be offered if tests are positive. To reach the aim of a 90% eradication rate with initial therapy, antibiotics should be tailored according to susceptibility testing. Therapy should be administered for 14 days, emphasizing strict adherence. Clarithromycin-containing regimens should be restricted to children infected with susceptible strains. When antibiotic susceptibility profiles are not known, high-dose triple therapy with proton pump inhibitor, amoxicillin, and metronidazole for 14 days or bismuth-based quadruple therapy is recommended. Success of therapy should be monitored after 4 to 8 weeks by reliable non-invasive tests [12].

The ESPGHAN/NASPGHAN guideline recommends many options for treating *H. pylori* if sensitivity is now known, including bismuth based regimen which was used in 5% of the studied population, high dose of amoxicillin in combination with metronidazole which was used in patients or concomitant therapy (PPI-Amoxicillin-Metronidazole-Clarithromycin) which was not used in the studied population. This outcome highlights the inadequate adherence of physicians to the ESPGHAN/NASPGHAN guideline, and necessitate increasing the awareness among treating physicians about the impotence of adhering to international recommendations.

Among the cohort of 345 (83.0%) patients who had documented adequate compliance and post-treatment eradication testing, the overall treatment success rate was 65%. Compared to the treatment success rate in early 1990s ranging from >80-90%, this data showed higher treatment failure rate, which is likely a reflection of high antibiotic resistance rates. Overall, the problem of antibiotics resistance is expanding globally and locally, in multiple bacterial pathogens [33], which is a critical issue requiring special attention. Within the developing world, treatment of *H. pylori* is hard due to the frequency of antibiotic resistance and high recurrence rates after successful treatment [5]. The antibiotic resistance of *H. pylori* strains in the UAE was studied by Al-Tayyari, he found that a significant proportion of gastric mucosal biopsies obtained in the UAE are positive for genes associated with clarithromycin and metronidazole resistance [29]. Similar findings were reported by Al-Faresi, among the 26 culture isolates, five 19.2% exhibited resistance to clarithromycin, while the remaining 24 were sensitive [30]. Recent studies highlight the need for tailored approaches due to varying eradication rates and antibiotic resistance.

The effectiveness of first-line therapy for *H. pylori* infection in children and adolescents varies depending on the treatment regimen used, which was the most significant predictor of treatment response in this study. This contradicts with some studies in adults reporting that eradication rates are significantly correlated with age (better in the middle age group), and non-smokers [34]. The latter study proposed that successful eradication of *H. pylori* infection is dependent on the combined effect of environmental, social, genetic, and clinical factors.

## Conclusion

In this retrospective study, the efficacy of first-line therapy for *H. pylori* was assessed in the pediatric and adolescent population of Abu Dhabi. The most effective first-line therapy was found to be PAC, which was the most widely used therapeutic regimen. These findings could be extrapolated to infer antibiotic resistance trends and potentially offer guidance for the broader region. The high prevalence of *H. pylori* in the study population is alarming, shedding light on the importance of screening tests and early eradiation of the bacteria to avoid chronic infection and subsequent future complications such as gastric cancers. To the best of our knowledge, there are no previous studies looking into the eradication rate of *H. pylori* therapy in the UAE and the Middle East. Thus, this study provides critical insights into the prevalence and success of treatment in our population, which is vital for proper public health action.

Decreasing eradication rates with previously recommended treatments calls for changes to first-line therapies and broader availability of culture or molecular-based testing to tailor treatment to the individual child [12]. More personalized approached may be required to ensure bacterial eradication. Novel therapeutics might be considered in the future to increase the success of treatment, such as microbiota-derived therapies like probiotics, phage therapy, *H. pylori* vaccine, natural products, and novel nanoparticles [35].

## Data Availability

Data cannot be shared publicly because of patients confidentiality

